# Exploring the role of generalized health biomarkers in Traumatic Brain Injury: A UK Biobank Study

**DOI:** 10.1101/2025.09.02.25334939

**Authors:** Shubhi Sharma, Donald M. Lyall

## Abstract

**Background:** The role of generalized health biomarkers remains widely unexplored in relation to TBI. However, they show association with worsened brain health and nervous system functioning. It is useful to explore potential relationship between TBI and generalized, circulating health biomarkers as surrogate endpoints that can be linked to severity and mortality.

**Aim:** This is a hypothesis-free study that investigates the potential relationship(s) between traumatic brain injury and 34 generalized, circulating health biomarkers using data collected by the UK Biobank.

**Methods:** Data from 419,778 white British participants are analyzed against 34 biomarkers from the UK Biobank database. TBI diagnosis is captured through ICD-10 codes and categorized as narrow-band and broad-band TBI. Multivariate linear regression analysis is conducted for changes in biomarkers in participants with TBI history vs. those with no history. This is analyzed across 4 models with increasing number of covariates and stratified for both categories of TBI.

**Results:** IGF-1, gamma glutamyltransferase and aspartase aminotransferase had significant results across all models in broad-band and narrow-band TBI (P < 0.05). An additional ten biomarkers showed statistically significant results in all models except the fully adjusted models.

**Conclusion:** IGF-1, GGT, and AST emerged as consistent indicators of systemic changes following TBI, highlighting their potential as surrogate markers for monitoring long-term outcomes. These findings suggest that incorporating generalized health biomarkers into clinical and research frameworks may improve early detection, risk stratification, and intervention strategies in TBI populations.

## Introduction

TBI is the leading cause for deaths and disabilities in adults under the age of 45 years, with over 69 million cases worldwide^1^. Despite its burden, TBI remains a poorly understood condition^2^. This is attributed to the condition’s heterogeneity, due its definition and severity remaining highly variable, and its impact on a wide range of granular physiological factors, as evidenced by patients with similar injuries experiencing distinct recovery trajectories and disease outcomes^3, 4^. It is appreciated that there are varied pathologies that may lead to physiological changes years before symptoms^5^; for example, hypopituitarism, Alzheimer’s disease and other dementia, neuroendocrine dysfunction and psychological disorders^6–9^. There is a need to explore post-TBI physiology to identify at-risk patients of Alzheimer’s disease.

Identifying and validating biomarkers that are significantly affected by TBI may be one way to reduce this gap^3^. Since the 1980s, biomarkers have been found to be valuable in health and medicine due to their cost effectiveness and measurability^3, 4^. However, current research focuses on TBI-specific biomarkers (like GFAP, S100B, NSE) that are not measured routinely because of the biomarkers’ they stringent requirement for specialized assays, equipment, and trained professionals^10^.

Generalized health biomarkers are used in routine health tests and provide insights about liver function, kidney function, lipid metabolism, glucose control, hormonal levels, and electrolyte balance^11^. Additionally, they are easily procured, and highly reproducible^11^. Various health biomarkers have an established relationship with TBI or, at least, with neurological health. As examples, vitamin D hormone (VDH), biologically active metabolite of vitamin D, has a protective role following TBI whereas its deficiency is hazardous^12^. IGF-1 may play a role in neuroprotective or neurorepair processes due to its upregulation on sites of TBI lesions^15^. Creatinine, potassium, sodium, and urea are associated with kidney function and essential for maintaining bodily homeostasis, and thus, may have a potential link to proper neurological health^16^.

UK Biobank is a cohort of nearly 0.5 million UK participants that attended baseline assessment across 12 centers between 2006 and 2010. These participants have detailed physical information as well as electronic health record linkage (EHR). We conducted hypothesis-free analyses to explore differences in generalized health biomarkers for people post-TBI, relative to no TBI.

## Methods

### Participants

Data were obtained from the UK Biobank which is a general population cohort with 502,490 participants. The data was collected between 2006 and 2010 from people between the ages of 40 and 69 years (mean = 56.5; SD = 8.1), across 22 assessment centers across the UK. Only participants of white British ancestry were examined because there is evidence of phenotypic and genetic complexity according to ancestry, and UKB participants are predominantly white British.

### Biomarkers

The UK Biobank collected data on 34 biomarkers (table 1).

**Table 1:**
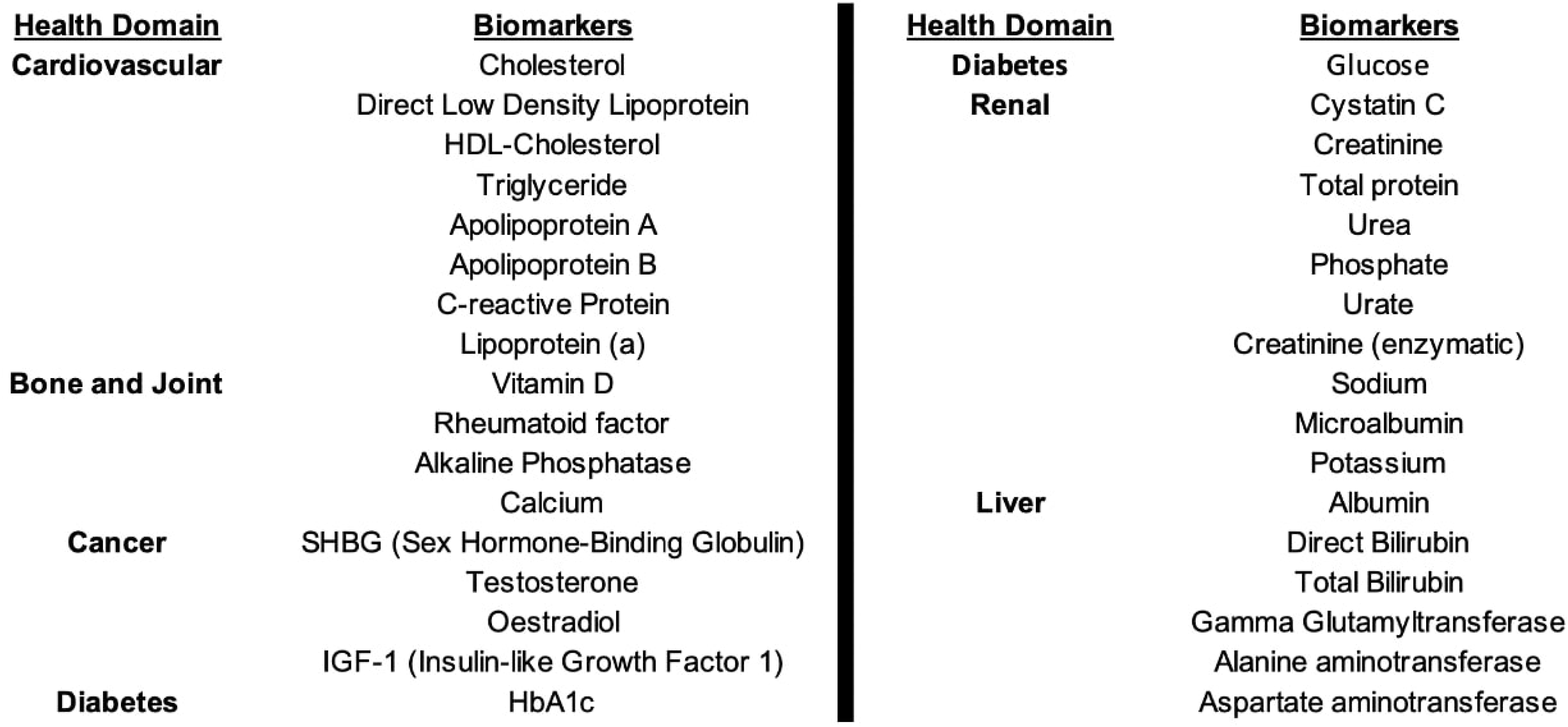
Generalized health biomarkers included in the UK Biobank inventory.

The biomarker data was collected at the initial appointment (as baseline) from all 502,412 participants and from 20,000 patients present at the follow-up assessment in 2012-2013. Quality control was strictly undertaken for measurements, instrumentation, and analyses^17^.

Oestradiol and rheumatoid factor (RF) were biomarkers with a relatively large number of values under measurable levels. Very low values were noted as ‘missing’ in the original data. However, if the participant reported significant values for a different biomarker, the oestradiol and/or RF value was recorded as the square root of the minimum agreed upon detectable value.

### TBI Classification

TBI diagnoses were based on self-report at baseline, or ICD-10 codes prior to that date (i.e., historic TBI). TBI diagnoses were stratified into narrow-band and broad-band TBI to reduce bias as a result of broad-band TBI cases that are less likely to be true TBI cases.

Traumatic brain injury diagnoses were recorded via linkage to primary or secondary diagnoses records using ICD-10 codes for hospital inpatient admission^18^. The diagnosis was classified into broad-band TBI, which included 1,798 ICD-10 codes related to head trauma, and narrow-band TBI, which was specific to TBI-related terminologies. Only TBI incidents prior to biomarker assessment were included. There was a total of 7,626 broad-band TBI cases, out of which 1,626 were narrow-band TBI cases.

### Study Design

This is a prospective, cross-sectional, cohort study.

Participants with TBI diagnosis have been assessed against biomarkers separately across four models with incremental adjustment:

1. <u>Model1</u> – unadjusted
2. <u>Model2</u> – partially adjusted for sex, age, BMI, and assessment centers.
3. <u>Model3</u> – intermediately adjusted for covariates from Model2, chronic neurological conditions (refer to table 2), and average sleep duration.
4. <u>Model4</u> – fully adjusted for covariates included in Model2 and Model3, Townsend deprivation indices, smoking history, educational qualification, heart disease, type 2 diabetes, high blood pressure, medication, and duration of moderate physical activity.

**Table 2:**
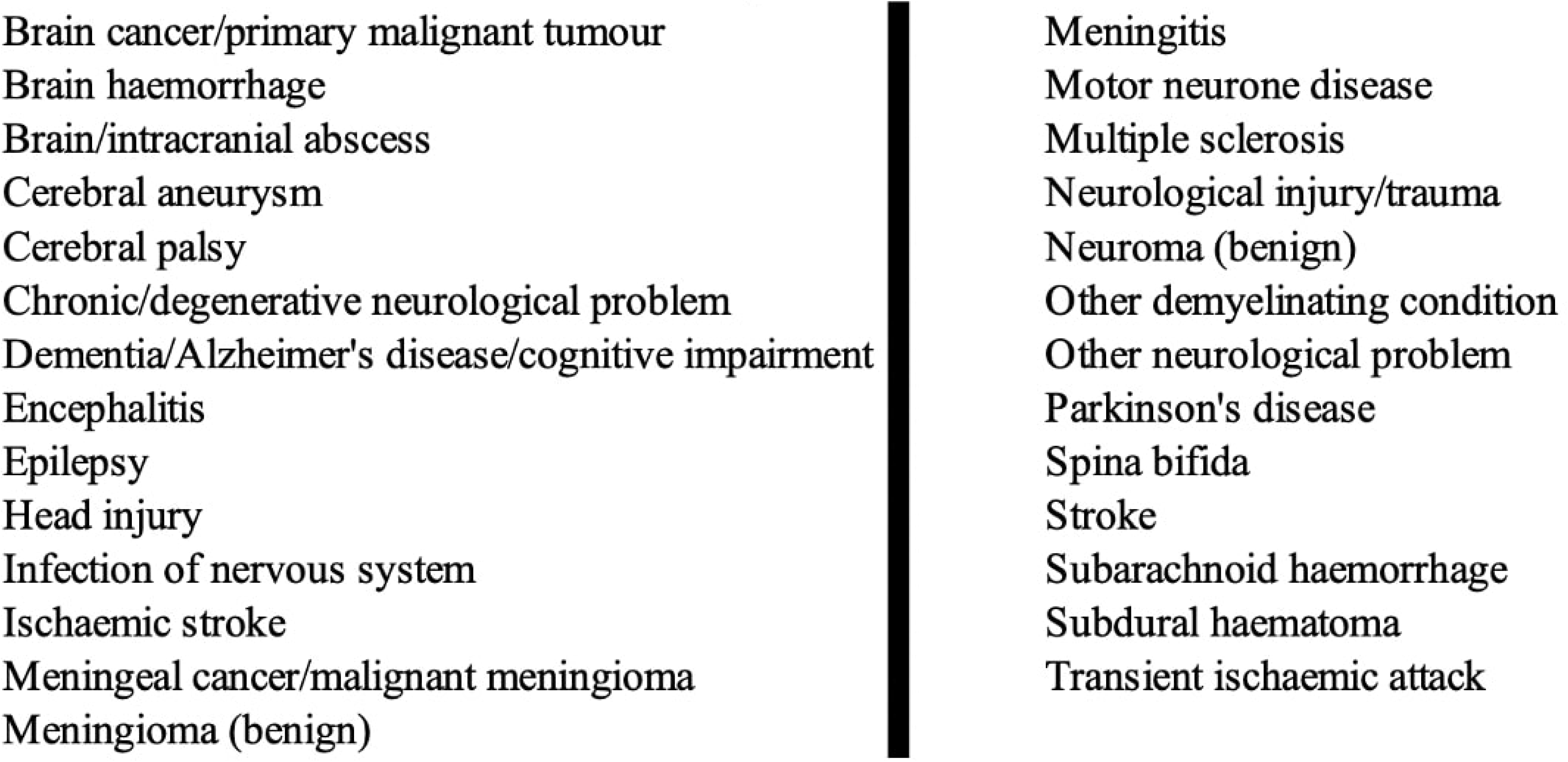
Chronic neurological conditions.

Using progressive models systematically would distinguish the specific impact of a confounding factor. Also, presenting multiple models would help in communicating transparency in the objective behind analyses. A fully adjusted model would account for the complexity in real world relationships and readily apply to a diverse demographic.

### Ethical Approval

Secondary data analysis is conducted using approval from the UK biobank ethics committee, NHS National Research Ethics Service and informed consent from all participants in the UK Biobank database for research, at the time of data collection.

### Data availability statement

UK Biobank is an open access resource available to affiliated researchers (http://www.ukbiobank.ac.uk/). Analysis syntax is available from the authors upon request.

### Covariates

Sex, age, weight, and height were measured during initial assessment visit. Assessment center refers to 1 of the 22 centers attended by participant. Educational qualification was self-reported and recorded as ‘1’ (at least university-level or college-level degree) or ‘0’. Duration of moderate physical activity was only accepted if response was between 0 and 1,440 minutes. Townsend deprivation indices were determined immediately prior to participation in the study based on postcode.

History of chronic neurological conditions (refer to table 2) was self-reported and included in the analyses as yes, i.e., history of at least 1 condition, vs. no. Sleep duration (in 24 hours) was self-reported. Smoking status was self-reported and collapsed into ‘never’ and ‘ever’, where ‘ever’ included both past and current smokers. Histories of heart disease, type 2 diabetes, and high blood pressure were self-reported and recorded individually. Self-report of medications was tabulated as yes vs. no.

### Statistical Analyses

Multivariate linear regression analysis was done using R version 2022.12.0+353. Results were expressed as standardized betas, defined as difference in the outcome variable in standard deviation units (e.g., ß = 0.2 would correspond to 0.2 SDs higher than average for TBI history vs. no TBI history). These are used to provide information about the strength and direction of the relationship. Corresponding P-values and adjusted R^2^ values were also calculated for each model across both strata.

## Results

The total number of participants, after exclusion, was 419,778; 7,626 are broad-band TBI, out of which 1,626 are narrow-band TBI. Distribution according to covariates can be found in tables 3 and 4.

**Table 3:** Summary of Descriptive Statistics.

| TABLE 3: Summary of Descriptive Statistics |  |  |  |  |
| --- | --- | --- | --- | --- |
| Covariates | No TBI History | TBI History |  | Missing Values |
|  |  | Broad-band | Narrow-band |  |
| N | 4,12,152 | 7,626 | 1,626 |  |
| Age (in years) | 56.7 ± 7.98 | 57.06 ± 8.32 | 56.29 ± 8.27 |  |
| Sex |  |  |  |  |
| Female | 187,216 (45.42%) | 3,901 (51.15%) | 989 (60.82%) |  |
| Male | 224,936 (54.58%) | 3,725 (48.85%) | 637 (39.18%) |  |
| Educational qualification | 128,052 (31.07%) | 2014 (26.41%) | 392 (24.11%) | 3,480 |
| BMI |  |  |  | 1,765 |
| <= 18.5 | 2,045 | 52 | 19 |  |
| > 18.5 and < 24.9 | 1,29,863 | 2,271 | 487 |  |
| >= 24.9 | 2,78,543 | 5,233 | 1,102 |  |
| History of chronic neurological condition | 17,158 (4.16%) | 882 (11.57%) | 292 (17.96%) | 16,532 |
| Avg. sleep duration (in hours) | 7.2 | 7.18 | 7.27 | 3,62,702 |
| Smoking status |  |  |  | 1,440 |
| Never | 225,143 (54.63%) | 3,775 (49.5%) | 753 (46.31%) |  |
| Ever | 185,601 (45.03%) | 3,819 (50.07%) | 863 (53.2%) |  |
| History of heart disease | 17,827 (4.33%) | 614 (8.05%) | 134 (8.24%) | 666 |
| History of type 2 diabetes | 19,050 (4.62%) | 546 (7.16%) | 127 (7.81%) | 928 |
| History of high blood pressure | 108,937 (26.43%) | 2,398 (31.45%) | 525 (32.29%) | 666 |
| Medication | 10,190 (2.47%) | 97 (1.27%) | 18 (1.11%) | 116 |
| Avg. moderate physical activity time (in min) | 66.7 | 73.54 | 76.94 | 1,09,662 |
| Avg. days b/w TBI diagnosis and assessment | N/A | 2,011 | 2,100 | 4,12,152 |
| Avg. townsend deprivation | -1.57 ± 2.92 | -0.71 ± 3.43 | -0.29 ± 3.61 | 507 |
| TOTAL = 419,778 |  |  |  |  |

**Table 4:** Frequency by Assessment Centre (in %)

| **TABLE 4: Frequency by Assessment**<br>**Centre (in %)** |  |  |  |
| --- | --- | --- | --- |
| **Centre**<br>**Code** | **No TBI**<br>**History** | **Broad-**<br>**band**<br>**TBI** | **Narrow-**<br>**band**<br>**TBI** |
| 10003 | 0.73 | 0.50 | 0.18 |
| 11001 | 2.63 | 2.02 | 2.40 |
| 11002 | 2.82 | 1.60 | 1.72 |
| 11003 | 3.67 | 7.03 | 2.28 |
| 11004 | 3.47 | 9.39 | 5.10 |
| 11005 | 3.36 | 9.31 | 3.14 |
| 11006 | 4.23 | 2.45 | 2.83 |
| 11007 | 6.02 | 3.21 | 4.00 |
| 11008 | 5.82 | 5.61 | 6.70 |
| 11009 | 8.01 | 7.02 | 6.89 |
| 11010 | 9.25 | 6.57 | 9.41 |
| 11011 | 9.11 | 6.91 | 8.36 |
| 11012 | 1.75 | 1.61 | 2.64 |
| 11013 | 7.13 | 5.57 | 6.70 |
| 11014 | 6.57 | 5.66 | 8.49 |
| 11016 | 6.77 | 8.81 | 10.76 |
| 11017 | 4.66 | 3.71 | 4.31 |
| 11018 | 4.38 | 3.32 | 3.75 |
| 11020 | 4.37 | 3.57 | 4.37 |
| 11021 | 4.64 | 4.16 | 5.35 |
| 11022 | 0.46 | 1.63 | 0.37 |
| 11023 | 0.14 | 0.34 | 0.25 |

P-Values for narrow-band TBI

Due to high missingness, average sleep duration and days between diagnosis and assessment were omitted from final analysis.

The results of the final analysis are reported in the form of standardized beta coefficients (ß), which are used to provide information about the strength and direction of the relationship between TBI history (independent variable) and the biomarker (dependent variable).

ß are presented through line plots in fig. 1 and fig. 2. The p-values heatmaps are intended to signify the analogous statistical significance (fig. 3 and fig. 4).

**Figure 1:**
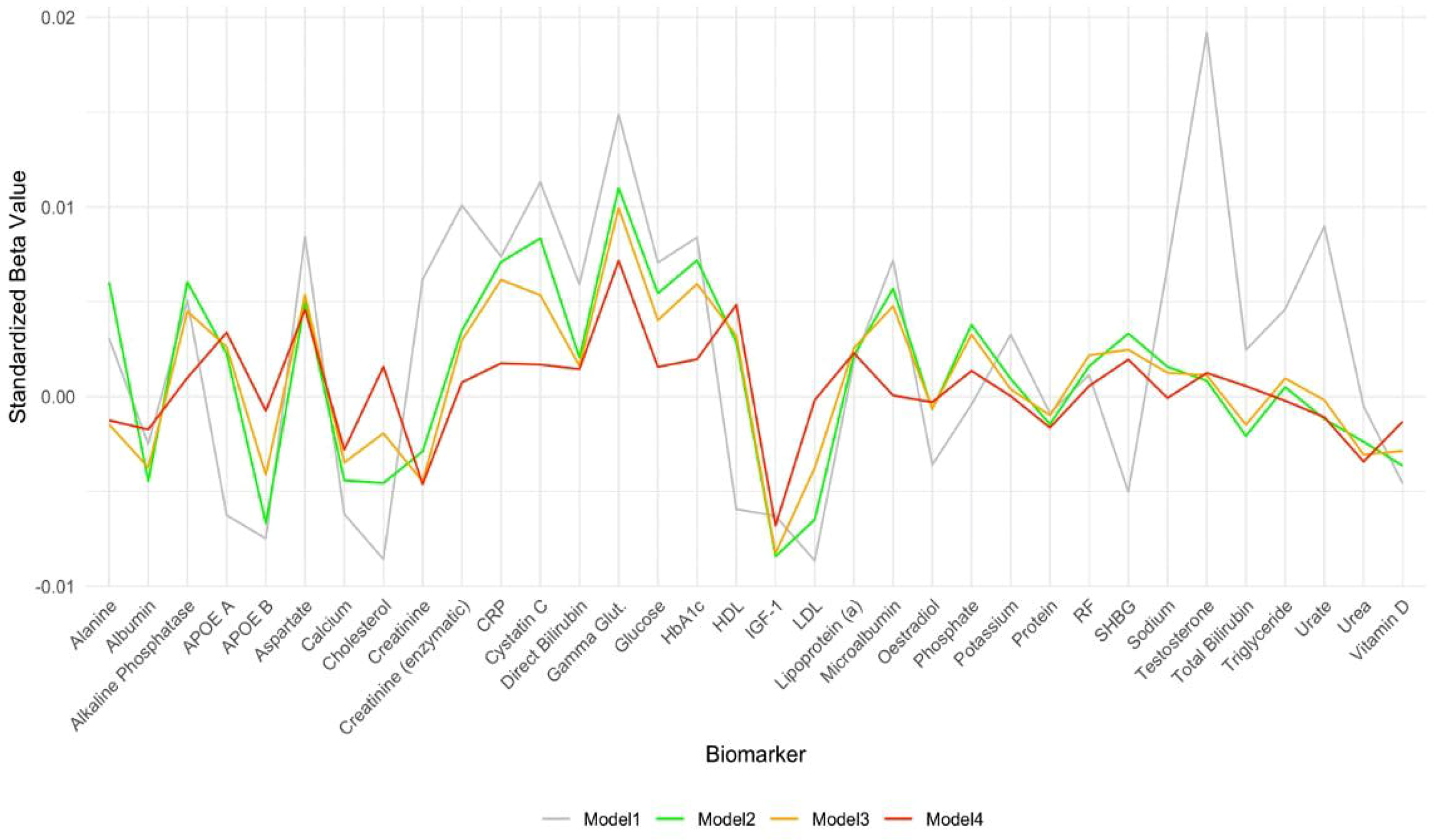
Line plot of standardized betas for narrow-band TBI.

**Figure 2:**
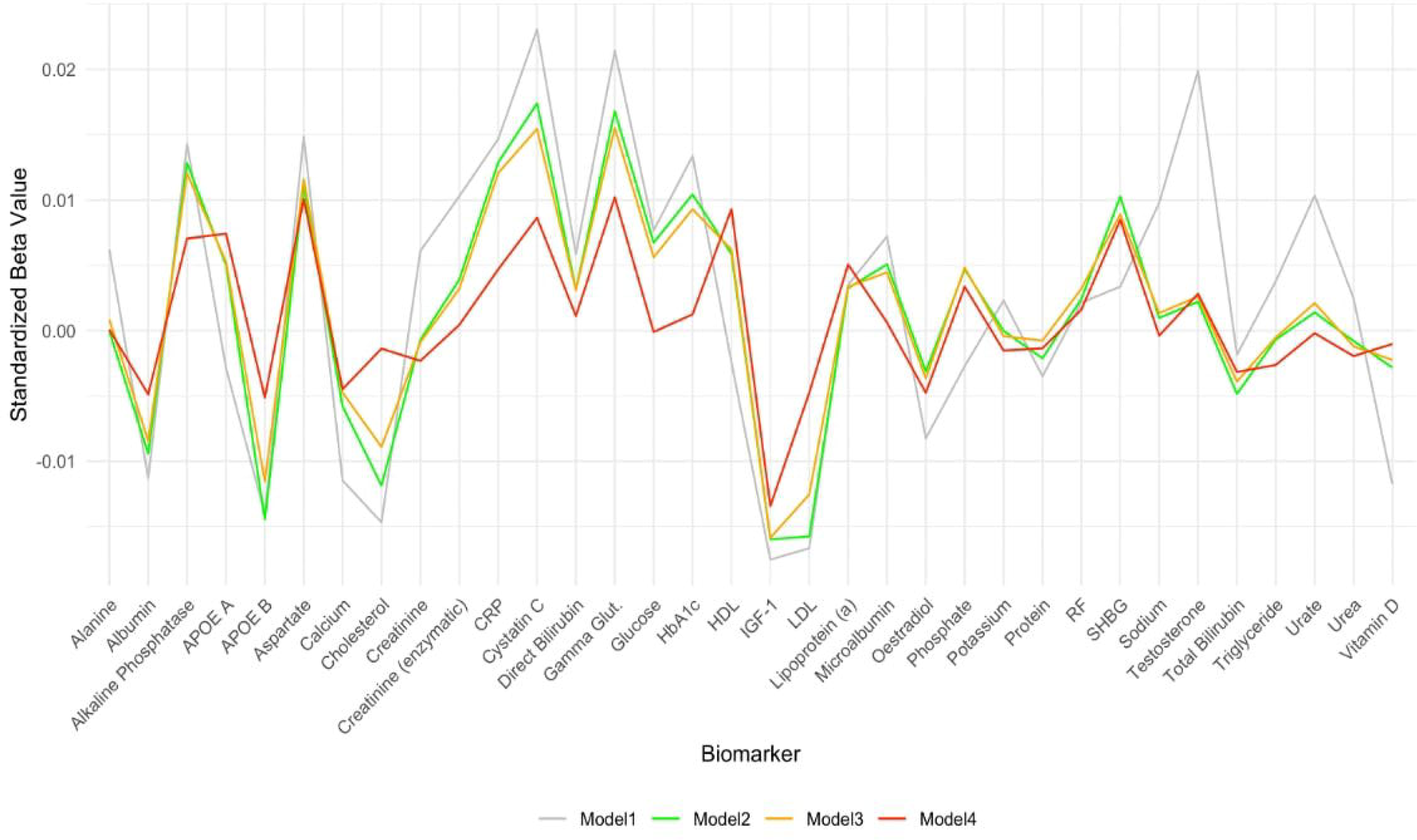
Line plot of standardized betas for broad-band TBI.

**Figure 3:**
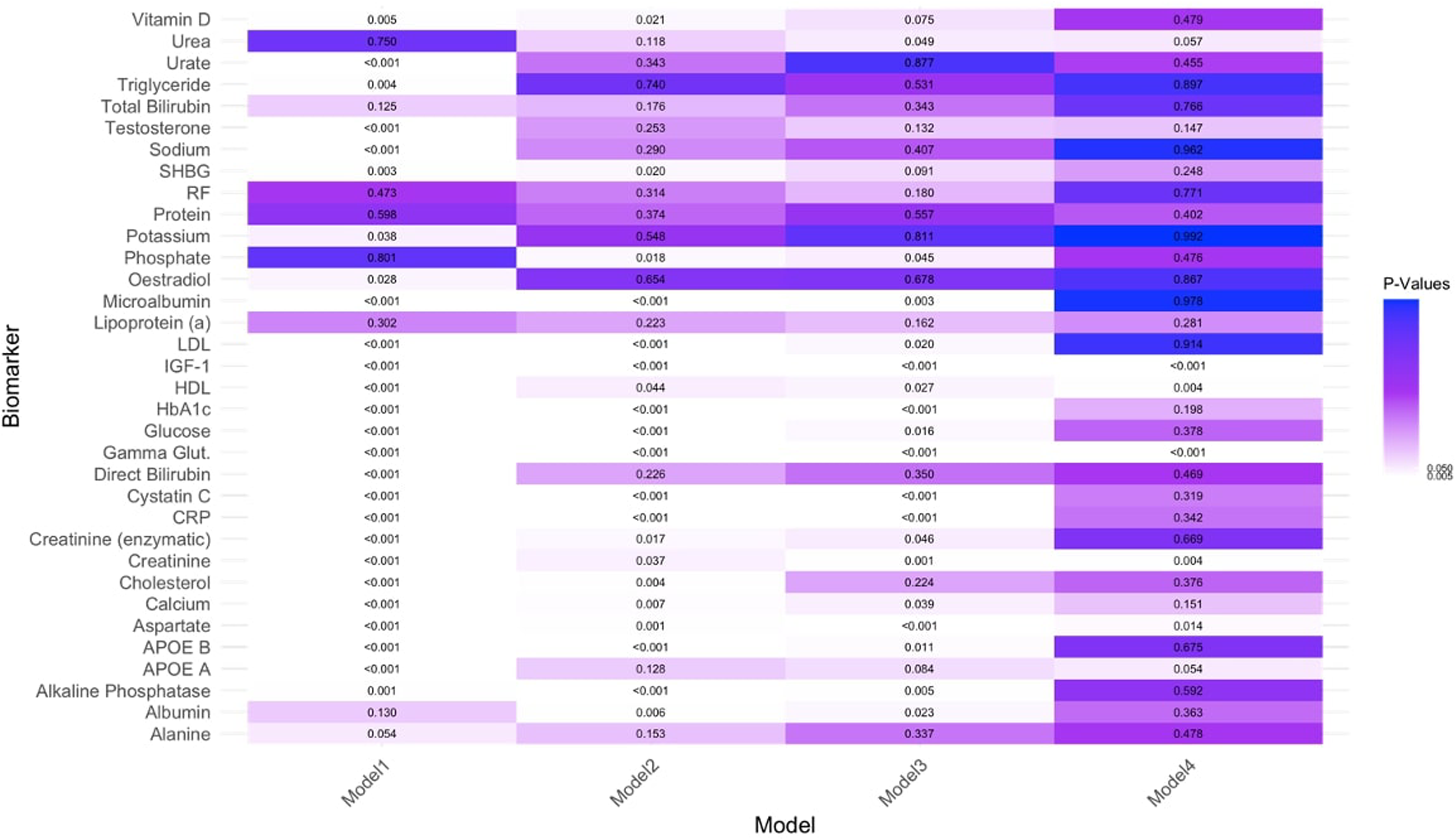
Heatmap of P-values for narrow-band TBI.

**Figure 4:**
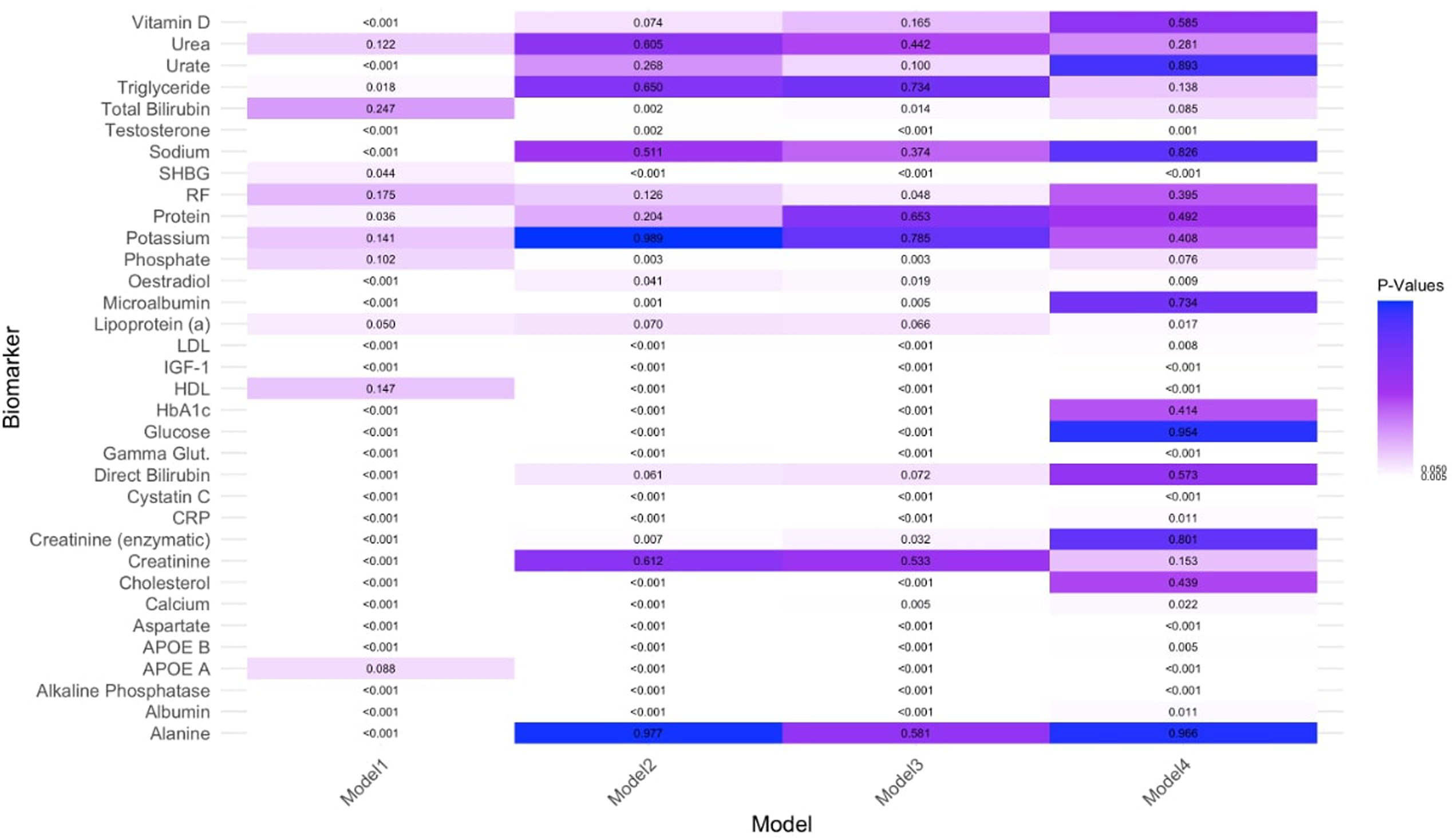
Heatmap of P-values for broad-band TBI.

The trends of statistical significance are evidenced in table 5, in which yellow boxes represent P < 0.05.

**Table 5:**
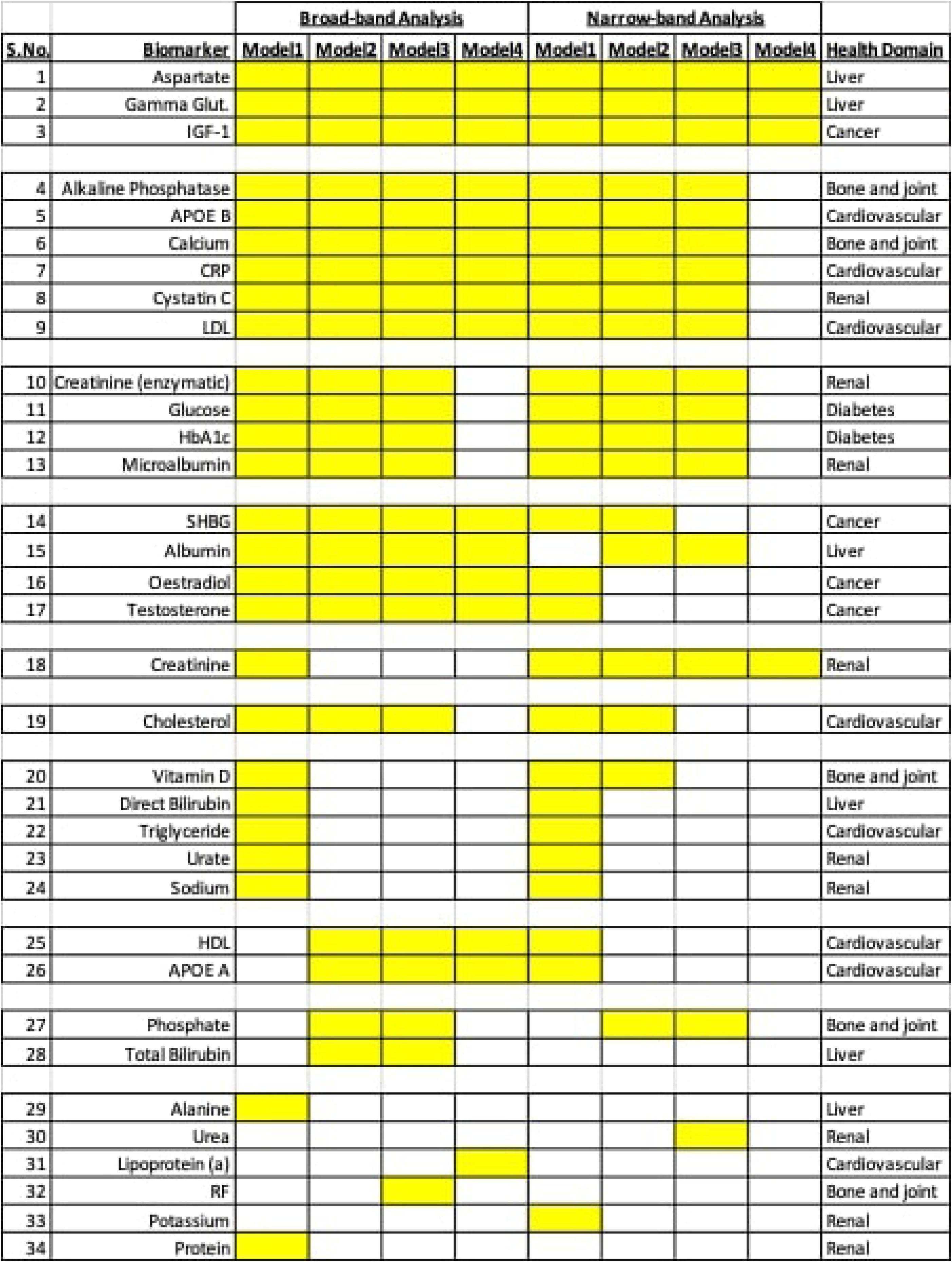
Summary of Significant Biomarkers.

Aspartate aminotransferase, gamma glutamyltransferase, and IGF-1 are statistically significant across all models in narrow-band and broad-band TBI. Alkline phosphatase, apolipoprotein A (APOE A), calcium, C-reactive protein (CRP), cystatin C, and direct low-density lipoprotein (LDL) are found to be statistically significant across all models, except Model4 of narrow-band TBI. Enzymatic creatinine, glucose, HbA1c, and microalbumin are statistically significant in all Model1, Model2 and Model3 of both narrow-band and broad-band TBI.

Sex hormone-binding globulin (SHBG), albumin, oestradiol, and testosterone have significant results in all models of broad-band TBI; for narrow-band TBI, only Model1 and Model2 have significant results for SHBG, only Model2 and Model3 for albumin and only Model1 in oestradiol and testosterone. Creatinine has significant results only in in Model for broad-band TBI but across all models in in narrow-band TBI.

For cholesterol, Model1, Model2 and Model3 showed significang results in broad-band TBI and Model1 and Model2 in narrow-band TBI. Vitamin D shows significant results in Model1 in broad-band TBI and Model1 and Model2 in narrow-band TBI. Direct bilirubin, triglyceride, urate and sodium have significant results in Model1 in both broad-band and narrow-band TBI.

HDL and APOE A has significant results in Model2, Model3 and Model4 in broad-band TBI and Model1 in narrow-band TBI. Phosphate has significant results in Model2 and Model3 in broad-band and narrow-band TBI. Total bilirubin has significant results in Model2 and Model3 in broad-band TBI.

Alanine aminotransferase, urea, lipoprotein (a), RF, potassium and total protein have one significant result across all 8 models of the analysis which is Model1 in broad-band, Model 3 in broad-band, Model4 in broad-band, Model3 in broad-band, Model1 in narrow-band and Model1 in broad-band TBI, respectively.

## Discussion

This study aimed to investigate the association between generalized health biomarkers and a history of traumatic brain injury (TBI) using data from the UK Biobank. Large-scale studies examining systemic health biomarkers in TBI are scarce, and this study provides new insights into how such biomarkers relate to both broad-band and narrow-band TBI cases.

The analysis identified three key biomarkers—insulin-like growth factor-1 (IGF-1), gamma-glutamyltransferase (GGT), and aspartate aminotransferase (AST)—that remained significant across all models in both broad-band and narrow-band TBI groups. These biomarkers suggest robust links between TBI and systemic physiological processes such as growth factor regulation, oxidative stress, and hepatic function. Furthermore, six additional biomarkers (apolipoprotein B, alkaline phosphatase, calcium, C-reactive protein [CRP], cystatin C, and low-density lipoprotein [LDL]) were significant in various models, particularly within narrow-band TBI cases. Conversely, four biomarkers (enzymatic creatinine, glucose, HbA1c, and microalbumin) lost statistical significance in the fully adjusted models, suggesting confounding influences from additional covariates.

### Interpretation of Significant Biomarkers

IGF-1 plays a fundamental role in neuroplasticity and neuronal repair. Previous research has demonstrated that IGF-1 is upregulated at sites of brain injury, promoting neuronal survival and synaptic plasticity^19^. Reduced serum IGF-1 levels in individuals with TBI may indicate impaired neuroprotective mechanisms, particularly in cases of chronic or repeated head trauma^20^. This aligns with animal studies where IGF-1 deficiency has been linked to hippocampal neuronal loss and cognitive decline post-TBI^21^. Additionally, low IGF-1 has been implicated in neurodegenerative diseases, further reinforcing its potential role in long-term TBI outcomes^22^.

Gamma-glutamyltransferase (GGT) is an enzyme involved in glutathione metabolism and oxidative stress regulation. Elevated GGT levels in TBI patients suggest increased oxidative stress, which has been linked to both neuroinflammation and blood-brain barrier disruption^23^. Oxidative stress is known to exacerbate neurodegeneration post-TBI, further highlighting the relevance of this biomarker^24^. Aspartate aminotransferase (AST) is traditionally used as a liver function marker but is also present in the brain. Elevated AST levels in TBI patients may reflect neuronal cell damage, similar to what has been observed in ischemic stroke cases^25^. Studies suggest that AST release into circulation may serve as an indirect marker of neuronal injury severity^26^.

### Additional Biomarkers and Their Implications

Increased ALP levels were observed in TBI patients, a finding previously associated with neurodegenerative diseases and systemic inflammation^27^. While its direct role in TBI remains unclear, ALP may be linked to neuroinflammatory pathways. Lower APO B levels suggest disruptions in lipid transport, which may impact neurovascular health and cognitive function in TBI patients ^28^.Hypocalcemia was noted in TBI cases, consistent with previous findings associating low calcium levels with poor neurological recovery and increased intracranial pressure^29^.Cystatin C, associated with kidney function and inflammation, was elevated in TBI patients. Higher cystatin C levels have been linked to neuroinflammatory conditions and cognitive impairment^30^.Elevated CRP levels indicate a strong inflammatory response following TBI. Chronic inflammation post-TBI has been associated with increased risk of neurodegenerative diseases such as Alzheimer’s and Parkinson’s disease^31^. Decreased LDL levels in TBI patients suggest disruptions in cholesterol metabolism, potentially affecting myelin integrity and neuronal function^32^.

### Comparing Narrow-Band vs. Broad-Band TBI Results

This study found stronger biomarker associations in the broad-band TBI group, likely due to its larger sample size and broader inclusion criteria. However, the narrow-band TBI group, consisting of more clinically confirmed cases, provided more targeted insights into TBI severity. The distinction between these two classifications is crucial, as mild and moderate TBI cases often go undiagnosed in hospital settings, leading to underrepresentation in broad-band datasets^33^. Differences in biomarker expression between these groups may reflect injury severity, underlying comorbidities, or post-injury systemic changes^34^.

### Clinical Implications and Future Research

1. Potential for Biomarkers as Surrogate Endpoints The findings suggest that generalized health biomarkers could serve as valuable surrogate markers for TBI prognosis. Given their routine use in clinical practice, these biomarkers could facilitate early identification of at-risk individuals and guide personalized intervention strategies. For instance, monitoring IGF-1 and GGT levels in TBI patients may provide insight into neuroinflammatory status and recovery trajectories^19^.
2. Feasibility and Practical Applications The accessibility of these biomarkers in routine health assessments underscores their potential for integration into clinical TBI management. Unlike specialized TBI-specific markers (e.g., GFAP, UCH-L1), which require advanced laboratory assays, the biomarkers identified in this study are widely measured in primary healthcare settings. This could enhance early diagnosis and long-term monitoring of TBI patients, particularly in resource-limited environments^35^.
3. Need for Longitudinal Studies Despite the strengths of this study, longitudinal research is necessary to establish causal relationships between these biomarkers and TBI progression. Future studies should investigate temporal changes in biomarker levels post-injury, exploring how fluctuations correlate with cognitive outcomes and rehabilitation efficacy. Additionally, the role of confounding factors, such as sleep disturbances and medication use, warrants further investigation^36^.

### Strengths and Limitations

This study uses a large sample size from the UK Biobank which enabled robust statistical analysis. This also allowed it to control for many demographic factors which could have confounding effects on the true relationship. External validity was preserved through stratification by TBI classification.

One of the biggest limitations of this study is the variability in biomarker collection methods which is further complicated by unreliability of non-fasting blood samples (which were used at various assessment centers). It is very poorly understood how biomarkers exit the nervous system to post-injury to enter the bloodstream^37^.

This study only includes participants of white British background and from a relatively less deprived background^40^. This reduces the external validity of results. Due to high rates of missingness, this study could not control for days between self-reported TBI diagnoses and assessment for biomarkers which has potential of being a significant confounding factor since its physiological effect is time dependent. This is evidenced by multiple previously cited studies. Sleep disturbances have been widely observed post-TBI^38^. This may also be a crucial confounding factor that wasn’t included in this study due to overwhelmingly large number of missing values.

## Conclusion

This study highlights the potential of generalized health biomarkers in understanding the systemic impact of TBI. The consistent associations observed for IGF-1, GGT, and AST suggest their role as key indicators of post-TBI physiological changes. While additional biomarkers exhibited significance in specific models, further validation is needed to explore their mechanistic underpinnings. The integration of these biomarkers into routine clinical practice could enhance diagnostic accuracy, improve patient monitoring, and facilitate early intervention strategies, ultimately contributing to better long-term outcomes for TBI patients.

