## Supplementary figures and images for "Exploring the role of generalized health biomarkers in Traumatic Brain Injury: A UK Biobank Study"

### Supplementary Table 1

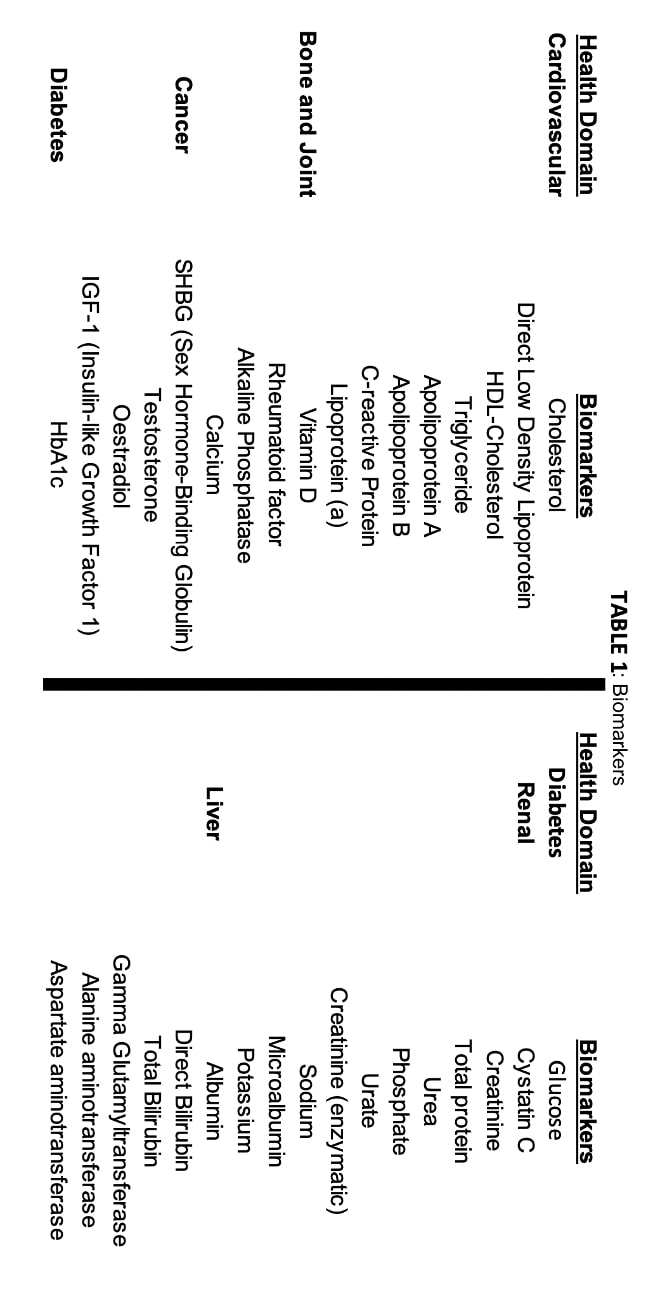

### Supplementary Table 2

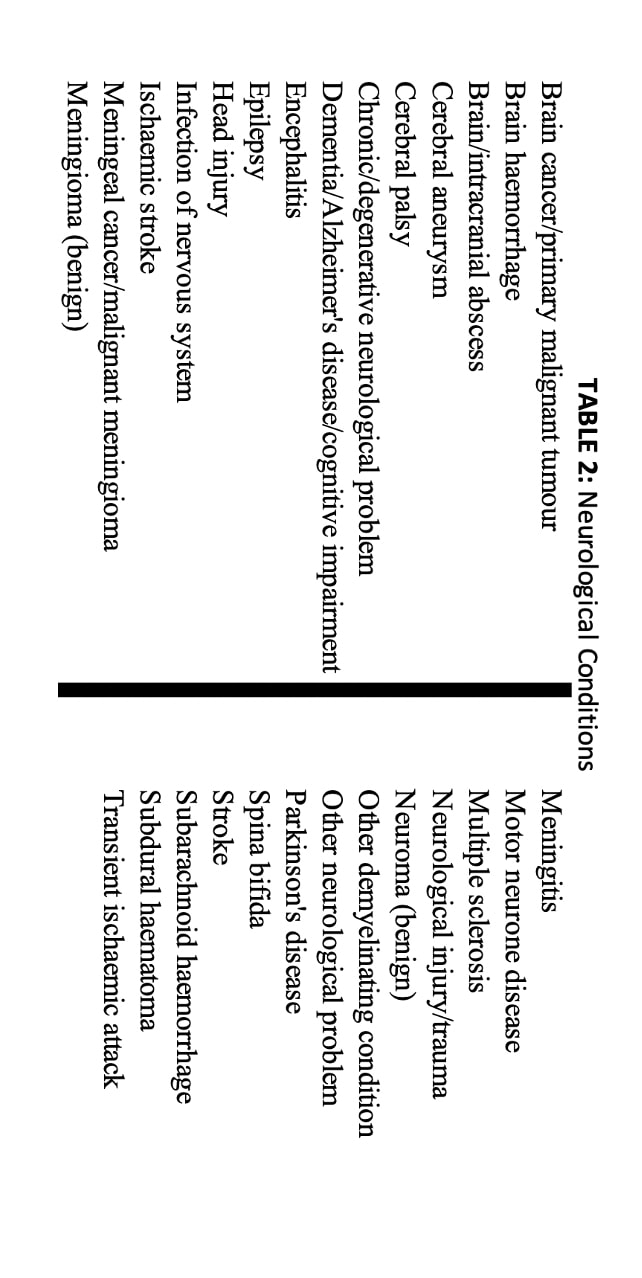

### Supplementary Table 3

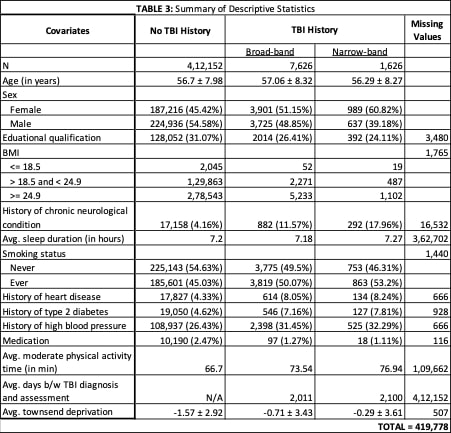

### Supplementary Table 4

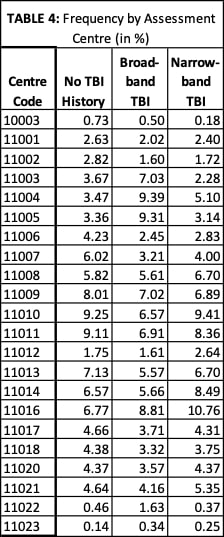

### Supplementary Table 5

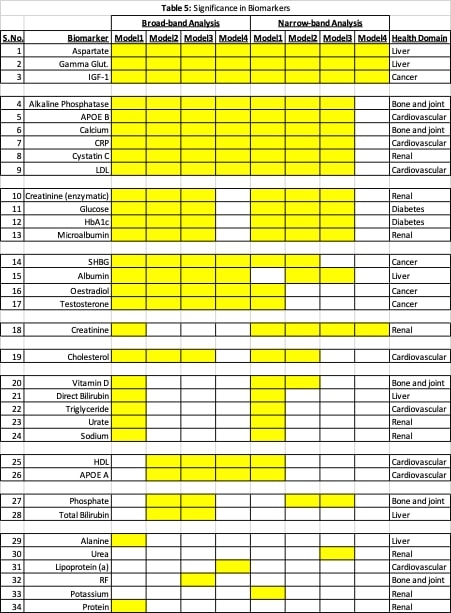
